# Clinical Data on Hospital Environmental Hygiene Monitoring and Medical Staff Protection during the Coronavirus Disease 2019 Outbreak

**DOI:** 10.1101/2020.02.25.20028043

**Authors:** Yanfang Jiang, Haifeng Wang, Yukun Chen, Jiaxue He, Liguo Chen, Yong Liu, Xinyuan Hu, Ang Li, Siwen Liu, Peng Zhang, Hongyan Zou, Shucheng Hua

## Abstract

**Background:** The outbreak of coronavirus disease 2019 (COVID-19) has placed unprecedented challenges on hospital environmental hygiene and medical staff protection. It is crucial to assess hospital environmental hygiene to understand the most important environmental issues for controlling the spread of COVID-19 in hospitals.

**Objective:** To detect the presence of COVID-19 in the samples from the area at risk of contamination in the First Hospital of Jilin University.

**Methods:** Viruses in the air were collected by natural sedimentation and air particle sampler methods. Predetermined environmental surfaces were sampled using swabs at seven o’clock in the morning before disinfection. The real-time reverse-transcription PCR method was used to detect the existence of COVID-19 pathogens.

**Results:** Viruses could be detected on the surfaces of the nurse station in the isolation area with suspected patients and in the air of the isolation ward with an intensive care patient.

**Conclusion:** Comprehensive monitoring of hospital environmental hygiene during pandemic outbreaks is conducive to the refinement of hospital infection control. It is of great significance to ensure the safety of medical treatment and the quality of hospital infection control through the monitoring of environmental hygiene.

## 1. Introduction

The outbreak of COVID-19 in Wuhan in December 2019 has led to a serious public health event ^[1-3]^. Meanwhile, the outbreak of this novel virus has placed unprecedented challenges on hospital environmental hygiene. The occurrence of medical staff-associated infections is closely related to long-lived pathogens in the hospital environment ^[4,5]^. Thus, it is crucial to assess hospital environmental hygiene to understand the most important environmental issues for controlling the spread of COVID-19 in hospitals. The Chinese government quickly adopted quarantine measures for confirmed and suspected patients to restrain the spread of the pandemic^[6]^. Comprehensive monitoring of hospital environmental hygiene during the outbreak of the pandemic is conducive to the refinement of hospital infection control ^[5,7]^. It also increases understanding of the environmental challenges corresponding to the reemergence of COVID-19 or similar viruses. Therefore, it is of great significance to ensure the safety of medical treatment and the quality of hospital infection control through the monitoring of environmental hygiene. According to a report from the China Centers for Disease Control and Prevention, as of February 11, 3019 medical staff members were infected with the new coronavirus (including confirmed cases, suspected cases, clinically diagnosed cases, and asymptomatic infections, of which 1716 were confirmed cases), indicating that infection through no occupational exposure may occur ^[8]^, but more evidence is needed to support this hypothesis. Meanwhile, it is extremely important to ensure the safety of medical treatments and the quality of hospital infection control by monitoring hospital environmental hygiene. However, sensitive and effective methods for monitoring hospital-acquired infection control are still limited. Here, by monitoring COVID-19 and detecting nucleic acid, we reveal clinical data indicating hospital environmental hygiene levels and provide a way to monitor pathogenic microorganism contamination and nosocomial infections.

## 2. Methods

### 2.1 Participant Characteristics

Fifteen suspected patients and one confirmed intensive care patient (ORF1ab and N were positive according to RT-PCR) were hospitalized in two isolation areas at the First Hospital of Jilin University, a unit for COVID-19 detection. The hospital carries out district management for these two types of patients, and each area includes a nursing station. We detected 158 samples of COVID-19 from the area at risk of contamination. This study was approved by the Ethics Committee of the First Hospital of Jilin University (Changchun, China).

### 2.2 Sampling and Sample Processing

The environmental monitoring methods referenced the hospital sanitation standards (GB15982-2012). All air was collected by two methods: natural sedimentation ^[9]^ and a microbial air sampler (MAS-100 ECO), for which the stream of air was set to exactly 100 liters/minute (Merck, Germany) ^[10]^. Environmental surfaces were sampled using swabs.

### 2.3 Primer and Probe Sequences

Two sequence regions (ORF1ab and N) that are highly conserved among sarbecoviruses were selected for primer and probe design. The primer and probe sequences for the ORF1ab gene assay were 5’-TGGGGYTTTACRGGTAACCT-3’ (forward; Y = C/T, R = A/G), 5’-AACRCGCTTAACAAAGCACTC-3’ (reverse; R = A/G) and 5’-TAGTTGTGATGCWATCATGACTAG-3’ (probe, in 5’-FAM/ZEN/3’-IBFQ format; W = A/T), and the primer and probe sequences for the N gene assay were 5’-TAATCAGACAAGGAACTGATTA-3’ (forward), 5’-CGAAGGTGTGACTTCCATG-3’ (reverse) and 5’-GCAAATTGTGCAATTTGCGG-3’ (probe, in 5’-FAM/ZEN/3’-IBFQ format). The expected amplicon sizes of the ORF1ab and N gene assays were 132 bp and 110 bp, respectively^[11]^. All primers and probes were purchased from a commercial source (Integrated DNA Technologies). The primer and probe sequences were subsequently confirmed to have perfect matches with other COVID-19 genome sequences available from the Global Initiative on Sharing All Influenza Data (GISAID; https://www.gisaid.org/; accession numbers: EPI_ISL_402119, EPI_ISL_402120, EPI_ISL_402121, EPI_ISL_402123 and EPI_ISL_402124; accessed 12 January 2020).

### 2.4 RNA Extraction

All the collected samples were inactivated by heating at 56 °C for 30 minutes. For RNA extraction from 600 µL of 0.9% NaCl solution in a 2.0 mL tube, the samples were centrifuged at 12000 g for 10 min and incubated for 30 min, the supernatant was discarded, 50 µL of RNA release agent was added to the tube, and the sample was mixed and incubated for 10 min.

### 2.5 Real-time Reverse-Transcription PCR

A 50-µL reaction contained 20 µL of RNA and 30 µL of 1× reaction buffer consisting of 26 µL of COVID-19 PCR mix (containing primers (4.62%), probes (1.15%), dNTPs (3.85%), MgCl^2^(0.77%), RNasin (0.48%), and PCR buffer (89.13%) (Shengxiang, Hunan, China)) and 4 µL each of COVID-19 PCR enzyme mix (containing RT enzyme (62.5%) and Taq enzyme (37.5%) (Shengxiang, Hunan, China)). Thermal cycling was performed at 50 °C for 30 min for reverse transcription, followed by 95 °C for 1 min and then 45 cycles of 95 °C for 15 s and 60 °C for 30 s. Participating laboratories used a Hongshi SLAN 96 S instrument (Hongshi, Shanghai, China). A sample was considered positive when the qPCR Ct value was ≤40.

## 3. Results

A total of 158 samples were collected from the area at risk of contamination. The two positive areas were the surfaces of the nurse station in the isolation area with suspected patients and the air of the isolation ward with an intensive care patient (Tables 2 and 3). We found that the virus was present both on surfaces and in the air. The total positive rate was 1.26% (2/158). The positive rates of the air and surface samples were 3.57% (1/28) and 0.77% (1/130), respectively.

**Table 1:** Collection of 158 samples from the area at risk of contamination

| Risk level | Classification | Sample collection points |  |
| --- | --- | --- | --- |
| High-risk area | CONTAMINATED AREA | Isolation ward area 1 (2F) | Isolation wards 1-15 door handle |
|  |  |  | Isolation wards 1-15 general subject surface |
|  |  |  | Isolation wards 1-15 indoor air |
|  |  |  | Isolation wards 1-15 window frames |
|  |  |  | Isolation wards 1-15 sides of a toilet handle |
|  |  |  | Nurse station general subject surface |
|  |  |  | Nurse station indoor air |
|  |  |  | Corridor exit general subject surface |
|  |  |  | General subject surface of the refuse storage area near the nurse station |
|  |  | Isolation ward area 2 (B1) | Isolation ward 16 door handle |
|  |  |  | Isolation ward 16 general subject surface |
|  |  |  | Isolation ward 16 indoor air |
|  |  |  | Isolation ward 16 window frames |
|  |  |  | Isolation ward 16 sides of a toilet handle |
|  |  |  | Nurse station indoor air |
|  |  | Fever clinic<br>2 consulting rooms | Door handle (inside and outside) |
|  |  |  | General subject surface |
|  |  |  | Indoor air |
|  |  |  | Computer keyboard |
|  |  | Fever clinic<br>observation room | Door handle (inside and outside) |
|  |  |  | General subject surface |
|  |  |  | Indoor air |
|  |  |  | Computer keyboard |
|  |  | Fever clinic<br>laboratory | Door handle (inside and outside) |
|  |  |  | General subject surface |
|  |  |  | Indoor air |
|  |  |  | Computer keyboard |
|  |  | Fever clinic<br>blood collection room | General subject surface |
|  |  |  | Indoor air |
| Medium-risk<br>area | SEMICONTAM<br>INATED AREA | Isolation ward area<br>buffer room | Door handle |
|  |  |  | General subject surface |
|  |  |  | Indoor air |
|  |  |  | Doctor's office window frames |
|  |  |  | Refuse storage area wall surface |
|  |  |  | Disinfection bucket surface |
|  |  |  | Dressing room ground |
|  |  |  | Medical staff of infection department<br>protective clothing |
|  |  | Pre-check triages | General subject surface |
|  |  |  | Infrared thermometer |
| Low-risk area | CLEAN AREA | Isolation ward area<br>working area | Entrance corridor general subject surface |
|  |  |  | Indoor air |
|  |  |  | Entrance corridor window frames |
|  |  |  | Physician's office door handle |
|  |  |  | Physician's office general subject surface |
|  |  |  | Director's office door handle |
|  |  |  | Door handle |

**Table 2:** Air monitoring results for the different risk areas

| Type |  | Negative | Positive | Positive rate |
| --- | --- | --- | --- | --- |
| High-risk area | Isolation ward | 19 | 1 | 5% |
|  | Fever clinic consulting rooms | 2 |  |  |
|  | Fever clinic observation room | 1 |  |  |
|  | Fever clinic laboratory | 1 |  |  |
|  | Fever clinic blood collection room | 1 |  |  |
| Middle-risk area | Isolation ward nurse station | 1 | 0 | 0 |
|  | Isolation ward buffer room | 1 | 0 | 0 |
| Low-risk area | Isolation ward clean area | 1 | 0 | 0 |
| Total |  | 27 | 1 | 3.57% |

**Table 3:**
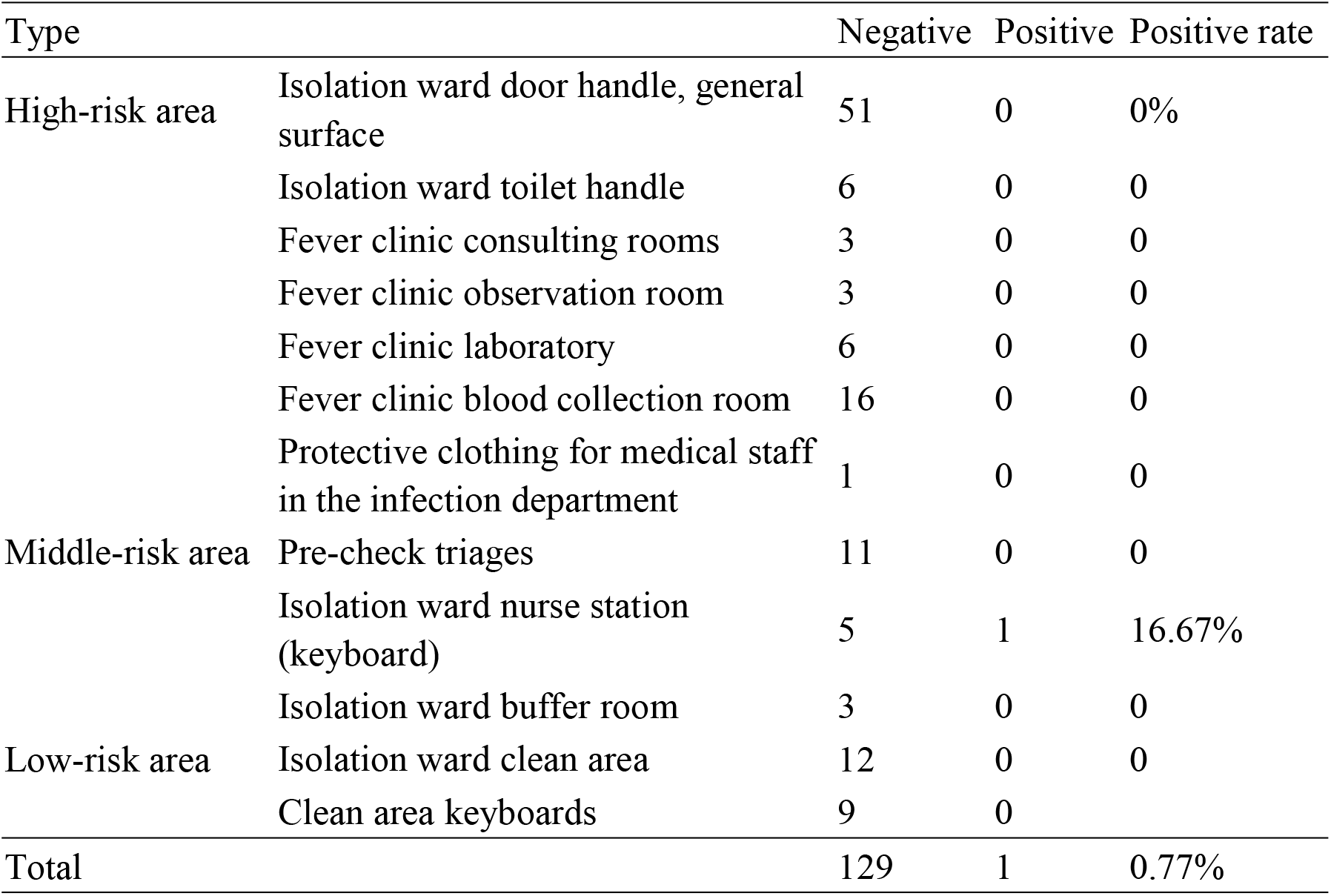
Surface monitoring results for the 154 different risk areas

Based on the original 24 hours of ultraviolet air filtering and 1000-2000 mg/L chlorine-containing disinfectant for ambient air and floor disinfection, the frequencies and duration times of air disinfections were extended. Key surfaces such as computer keyboards that were easily overlooked were clearly noted and carefully disinfected. The samples from the positive area and indoor air were collected 24 hours later, and the test results were negative.

## 4. Discussion

Our positive results in nucleic acid tests indicated that viruses were present in the air of an isolation ward with intensive care patients. The hermetic spaces of isolation wards are weak in air flow, which may cause high contact with the virus. Additionally, the confirmed patient underwent tracheal intubation the day before the samples were collected, and this procedure produced large amounts of aerosols that spread in the wards and seriously polluted the air. Siegel et al. reported that medical ventilators might generate respiratory aerosols that have been associated with an increased risk of occupationally acquired infections among healthcare personnel ^[12]^. Therefore, it is necessary to increase the intensity of disinfection for isolation wards with intensive care patients. For example, we should enhance ventilator exhaust management by adding ventilator exhaust port filters.

The suspected and confirmed patients stay a relatively long time in the high-risk area and may influence the environment. The intrusive and other operations that can easily produce aerosols may cause serious air pollution. The sites under observation without patients and operations have less impact on the environment. The samples from the same collection points were tested after a series of infection prevention and control measures were taken, such as continuous use of an air disinfection machine for the disinfection of ward air; extending the frequency of disinfection for ground and object surfaces; changing gloves and cleaning hands after operations and leaving the ward; and covering computer keyboards and changing the cover each day. Following the implementation of these measures, the results were negative, showing the effectiveness of disinfection. In the high-risk areas, such as hospital pre-check triages and the fever clinic, due to the open environment and large amount of air flow, the results were negative.

The previous research reports that the routine cleaning and disinfection procedures are the most likely cause of the deficiency of disinfection in the hospital environment, leading to the spread of pathogens ^[13]^. In our study, viruses were detected on the surfaces of the nurse station in the isolation area with suspected patients, suggesting that we need to strengthen the surface disinfection of nurse stations, especially focusing on computer keyboards, mice and types of equipment that are not easy to disinfect. A large number of studies have shown that the hands or gloves of medical staff members may be contaminated by contact with environmental surfaces contaminated by pathogens^[13-15]^. In the process of diagnosis and treatment, pathogens may be transmitted to patients and cause hospital-associated infections. Therefore, it is also very important to further strengthen the hand hygiene of the medical staff. We need to formulate refinement measures based on environmental hygiene monitoring data to improve the quality of hospital infection control. Due to fact that “uncultivable” microbes widely exist, traditional methods used to measure the effect of hospital infection control, such as sedimentation (exposing a microbial growth plate to the environment) have shown limited effects and poor sensitivity. For laboratories and testing organizations, high concentrations of nucleic acid may exist in aerosols, which may influence the test results and the operator’s safety and may even cause spread of the disease. Nucleic acid detection provides an effective method with which to monitor the environment and to evaluate the effectiveness of disinfection, especially for highly contagious diseases or pathogenic microorganisms with a potential aerosol risk, such as COVID-19. These nucleic acid detection results are very important for standardized hospital infection control. This study also increases our understanding of the environmental challenges corresponding to the reemergence of COVID-19 or similar viruses.

The data obtained in this study suggest that COVID-19 exists in the air of isolation wards with intensive care patients and existed on the surfaces of nurse stations and that medical staff encounter this virus in the air and on surfaces during patient care activities. An effective disinfection procedure may reduce the biosafety risk.

For hospital infection control, we suggest:

1. Different people should be responsible for the isolation observation area and the isolation ward area; crossing the areas should be strictly prohibited.
2. Objects in each area should be used exclusively; equipment that must be used for special purposes should be sterilized before being used in other areas.
3. Hand hygiene rules should be observed strictly, thoroughly and at all times.
4. Additional gloves, disposable isolators, protective screens or hoods should be worn when an operation may cause the spattering of blood, bodily fluid or aerosol, and these items should be discarded immediately to avoid the pollution of other areas.

## Data Availability

All data, models, and code generated or used during the study appear in the submitted article.

## Acknowledgments

This work was supported by the National Natural Science Foundation of China (Nos. 30972610 and 81273240), the Jilin Province Science and Technology Agency (Nos. 20160101037JC, 20170622009JC, 2017C021, and 2017J039), the Norman Bethune Program of Jilin University (2012206), and Open Project Funding From the State Key Laboratory of Kidney Diseases. We thank the patients and their families for participating in this study. We would also like to acknowledge the professional manuscript services of American Journal Experts.

## Disclosure Statement

The authors have no conflicts of interest to declare.

## Notes

### Competing Interest Statement

The authors have declared no competing interest.

### Funding Statement

the National Natural Science Foundation of China (Nos. 30972610 and 81273240), Jilin Province Science and Technology Agency (Nos. 20160101037JC, 20170622009JC, 2017C021, and 2017J039), Norman Bethune Program of Jilin University (2012206), and Open Project Funding from State Key Laboratory of Kidney Diseases.

## References

1. Benvenuto, D., et al., The global spread of 2019-nCoV: a molecular evolutionary analysis. Pathog Glob Health. 2020, 12: 1–4.

2. Huang, C., et al., Clinical features of patients infected with 2019 novel coronavirus in Wuhan, China. Lancet. 2020, 395(10223): 497–506.

3. The, L., Emerging understandings of 2019-nCoV. Lancet. 2020, 395(10221): 311.

4. Anderson, D.J., et al., Enhanced terminal room disinfection and acquisition and infection caused by multidrug-resistant organisms and Clostridium difficile (the Benefits of Enhanced Terminal Room Disinfection study): a cluster-randomised, multicentre, crossover study. Lancet. 2017, 389(10071): 805–814.

5. Beggs, C., et al., Environmental contamination and hospital-acquired infection: factors that are easily overlooked. Indoor air. 2015, 25(5): 462–74.

6. Wang, D., et al., Clinical Characteristics of 138 Hospitalized Patients With 2019 Novel Coronavirus-Infected Pneumonia in Wuhan, China. JAMA. 2020.

7. Phan, L.T., et al., Respiratory viruses in the patient environment. Infect Control Hosp Epidemiol. 2020, 41(3):259–266.

8. Chinese Center for Disease Control and Prevention, Beijing 102206, China. Epidemiology group of new coronavirus pneumonia emergency response mechanism, Chinese center for disease control and prevention, The epidemiological characteristics of an outbreak of 2019 novel coronavirus diseases (COVID-19) in China, Zhonghua Liu Xing Bing Xue Za Zhi. 2020, 41(2):145–151.

9. Odebode A, et al., Airborne fungi spores distribution in various locations in Lagos, Nigeria.Environ Monit Assess. 2020, 192(2):87.

10. Irie K, et al., Investigation of the detection ability of an intrinsic fluorescence-based bioaerosol detection system for heat-stressed bacteria.PDA J Pharm Sci Technol. 2014, 68(5):478–93.

11. Chu DKW,et al., Molecular Diagnosis of a Novel Coronavirus (2019-nCoV) Causing an Outbreak of Pneumonia. et al., Clin Chem. 2020, pii: hvaa029. doi: 10.1093/clinchem/hvaa029.

12. Siegel, J.D., et al., 2007 Guideline for Isolation Precautions: Preventing Transmission of Infectious Agents in Health Care Settings. Am J Infect Control. 2007, 35(10 Suppl 2): S65-164.

13. Pirincci, E. and B. Altun, An analysis of hospital cleaning staff’s attitudes and conduct regarding hand hygiene and cleaning. Int J Occup Saf Ergon. 2016, 22(2): 241–5.

14. Pittet, D., et al., Evidence-based model for hand transmission during patient care and the role of improved practices. Lancet Infect Dis. 2006, 6(10): 641–52.

15. Christensen, T.E., J.S. Jorgensen, and H.J. Kolmos, [The importance of hygiene for hospital infections]. Ugeskr Laeger. 2007, 169(49): 4249–51.

